# An LDLR missense variant poses high risk of familial hypercholesterolemia in 30% of Greenlanders and offers potential for early cardiovascular disease intervention

**DOI:** 10.1101/2022.01.12.22269132

**Authors:** Emil Jørsboe, Mette K. Andersen, Line Skotte, Frederik Filip Stæger, Nils J. Færgeman, Kristian Hanghøj, Cindy G. Santander, Ninna Karsbæk Senftleber, Lars J. Diaz, Maria Overvad, Ryan K. Waples, Frank Geller, Peter Bjerregaard, Mads Melbye, Christina V. L. Larsen, Bjarke Feenstra, Anders Koch, Marit E. Jørgensen, Niels Grarup, Ida Moltke, Anders Albrechtsen, Torben Hansen

## Abstract

**Background:** The common Arctic specific *LDLR* p.G137S variant was recently shown to be associated with elevated lipid levels. Motivated by this we aimed to investigate the effect of p.G137S on metabolic health, and cardiovascular disease risk among Greenlanders to quantify its impact on the population.

**Methods:** In a population based Greenlandic cohort (n=5063), we tested for associations between the p.G137S variant and metabolic health traits as well as cardiovascular disease risk based on registry data. Additionally, we explored the variant’s impact on **plasma NMR measured** lipoprotein concentration and composition in another Greenlandic cohort (n=1629).

**Results:** 29.5% of the individuals in the cohort carried at least one copy of the p.G137S risk allele. Furthermore, 25.4% of the heterozygous and 54.7% of the homozygous carriers had high levels (>4.9 mmol/L) of LDL cholesterol, which is above the diagnostic level for familial hypercholesterolemia (FH). Moreover, p.G137S was associated with an overall atherosclerotic lipid profile, and increased risk of ischaemic heart disease (HR (95% CI), 1.51 (1.18-1.92), P=0.00096), peripheral artery disease (1.69 (1.01-2.82), P=0.046), and coronary operations (1.78 (1.21-2.62), P=0.0035).

**Conclusions:** Due to its high frequency and large effect sizes, p.G137S has a marked population-level impact, increasing the risk of FH and cardiovascular disease for up to 30% of the Greenlandic population. Thus, p.G137S is a potential marker for early intervention in Arctic populations.

## Introduction

Cardiovascular disease (CVD) is the number one cause of death in many populations worldwide ^1^. Also among Greenlanders, the CVD burden is high, and the CVD prevalence is likely to increase in the future, due to increasing life expectancy and changing lifestyle ^2,3^. Therefore, additional focus on prevention and treatment of CVD is essential, also in the Greenlandic population. CVD is tightly linked to alterations in the lipid homeostasis, including elevated levels of LDL cholesterol ^4–7^. Alterations in the lipid homeostasis is associated with lifestyle factors such as unhealthy diet, physical inactivity, smoking, and alcohol intake, as well as genetic predisposition. The genetic predisposition can be monogenic, where a single variant has a large effect on lipid levels causing familial hypercholesterolemia (FH) ^8^, or polygenic where a large number of common variants each contribute with a small increase in the risk of dyslipidemia. The monogenic forms of dyslipidemia are associated with increased risk of CVD, especially ischemic heart disease ^9^. And on average, individuals with FH die 19 years earlier than the general population from a CVD event ^10,11^. Importantly, monogenic FH mutations seem to impose a two-fold higher risk of CVD, compared to higher LDL cholesterol caused by polygenic variants, likely due to the fact that monogenic variants manifest earlier in life than polygenic variants, leading to a greater cumulative LDL cholesterol exposure ^12,13^. Early treatment with cholesterol-lowering drugs has been shown to improve health outcomes for FH patients, and current guidelines recommend cholesterol-lowering treatment beginning from the age of 8 to 10 years ^9,14,15^. Hence, it is of great importance to identify and treat individuals carrying FH causing mutations early in life to reduce morbidity and mortality. The most common cause of FH is mutations in the gene (*LDLR*) encoding the LDL receptor (LDLR). This cell surface receptor mediates the uptake of LDL-cholesterol particles from the blood, primarily into the liver. The hepatic LDL uptake suppresses de novo synthesis of cholesterol, further reducing the concentration of circulating cholesterol ^16^.

In Arctic populations, a common *LDLR* missense variant (p.G137S; rs730882082) has been identified. This variant is located in exon 4, which encodes the ligand binding domain of the receptor, and has been predicted to be deleterious ^17^, and to reduce LDLR ligand binding by around 60%^18^. The p.G137S variant has been shown to be associated with markedly elevated levels of LDL cholesterol, total cholesterol, and apolipoprotein B ^18^, yet it is not known if the variant is associated with increased risk of CVD. Motivated by this, we aimed to investigate the influence of the common Arctic specific *LDLR* p.G137S missense variant on the metabolic and cardiovascular health in Greenlanders.

## Methods

### Study Cohorts

The cohort used in all analyses, except for the NMR analysis, is comprised of Greenlanders living in Greenland from The Population Study in Greenland 1999 (B99; n = 1401) and The Inuit Health in Transition Study (IHIT; n = 3115), as well as Greenlanders living in Denmark (BBH; n = 547), all collected as a part of general population health surveys of the Greenlandic population, which took place in 1999-2001 and 2005-2010 ^19,20^. Altogether, 295 individuals participated in both B99 and IHIT, and were treated as a separate survey when adjusting for survey in the statistical analyses. For the NMR analysis, we used data from a separate Greenlandic cohort collected in 2013 as part of a population-based sample (n = 1629), as previously described ^21^.

### Genetic data

The *LDLR* p.G137S variant (rs730882082) was genotyped using the KASP Genotyping Assay (LGC Genomics). The genotyping call rate was 99.42%, and 0 mismatches were observed in 361 samples genotyped in duplicate. We used previously published genome-wide genotype data from the same individuals to obtain estimates of a genetic similarity matrix needed in the association analyses. This genome-wide genotype data was generated using the Illumina MetaboChip (Illumina, San Diego, CA, USA, ^22^) and consisted of data from 4674 of the Greenlandic participants (IHIT, 2791; B99, 1336; BBH, 547) and 115,182 SNPs after quality control. Both the genotyping procedure and quality control for the SNP chip dataset have previously been described in detail ^23^. For the *LDLR* p.G137S variant genotyping and the MetaboChip data there was an overlap of 4653 individuals. This is the data set we based our analyses of metabolic phenotypes on.

For the cohort with NMR data, the genome-wide genotype data was generated using the Illumina OmniExpressExome SNP chip (Illumina, San Diego, CA, USA). After quality control this cohort comprised 1570 participants and genotypes of 643,734 SNPs ^21^.

### Analysis of metabolic phenotypes

Since the Greenlandic study cohorts contain admixed individuals with both Inuit and European ancestry, and numerous close relatives ^24^, we used a linear mixed model to perform the association analyses. This type of model takes admixture and relatedness into account by including them as a random effect. Specifically, we used the linear mixed model software tool GEMMA (v0.95alpha) ^25^ for quantitative traits and GMMAT ^26^ for binary traits. To perform the analyses we first estimated a genetic similarity matrix for all participants with both genotype and phenotype data available from quality controlled genome-wide SNP chip data. The estimation was performed by applying GEMMA to standardised genotypes from the SNPs with MAF of at least 5% and missingness of maximum 1%. Association tests were then performed using a score test in GEMMA and a Wald test in GMMAT and effect sizes and standard errors were estimated using a restricted maximum likelihood (REML) approach. For the quantitative traits we used a rank-based inverse normal transformation prior to analysis and we reported effect size estimates in standard deviations *β_SD_*. This transformation was done independently for men and women. To get effect size estimates in the measured units, we also performed analyses of non-transformed trait values. In all tests we assumed an additive model and included sex, age and survey (IHIT, B99, BBH) as covariates.

### Analysis of CVD data

We used the R-package survival ^27^ for Cox regression analyses, adjusting for sex, age, survey, and the top 10 genetic principal components (PCs), based on the genome-wide genotype data. The top 10 PCs were included to correct for population structure. Follow-up time was calculated as years lived from birth to an event, death of other causes, emigration, or end of follow-up (December 31th 2016). A second analysis was done in the same way, but with follow-up time calculated as years from inclusion in study, and thereby excluding individuals with a CVD event of that type before the study initiation. Also a logistic regression using GMMAT, in the same way as previously described, was done for each type of CVD event defining cases as anyone with a CVD event of that type, to estimate the cross-sectional CVD risk.

More detailed information on the methods used is available in the supplementary material.

## Results

In the Greenlandic population, which has both Inuit and European ancestry, we estimated that the *LDLR* missense variant (p.G137S) had a minor allele frequency (MAF) of 15.8%, and that the minor allele was carried by 29.5% of the population. Within the Inuit ancestry component of the population we estimated a MAF of 22.7% and of 0.0% in the European ancestry component. In line with this we estimated the variant to be extremely rare (MAF<0.005%) in non-Arctic populations (Table S1). Even though p.G137S is a missense variant it might still act as an expression quantitative trait locus however, we observed no association between p.G137S genotype and expression of *LDLR* RNA in blood (P = 0.22; Figure S1).

### Metabolic phenotypes

We first assessed how the p.G137S variant affected a range of metabolic traits in a cohort of 4653 Greenlanders. We confirmed a strong association between the A-allele of the p.G137S variant and elevated concentrations of circulating LDL cholesterol (β, 0.75 mmol/L, P=5.5·10^−87^, Table 1 and Figure 1A). The effect was additive, and similar across age groups (Figure S2), as well as degree of Inuit ancestry (Figure S3). As shown in Figure 1B, we observed that 8.0%, 25.4%, and 54.7% of non-, heterozygous, and homozygous p.G137S A-allele carriers, respectively, had high LDL cholesterol levels (>4.9 mmol/L), which is the diagnostic threshold for familial hypercholesterolemia ^9^. We noticed an even larger effect of the variant on LDL cholesterol levels, when also including individuals prescribed cholesterol-lowering drugs, especially among homozygous carriers (Table 1 and Figure S4). Of note, prescription of cholesterol-lowering drugs was significantly associated with higher LDL cholesterol levels, across all individuals, as well as only heterozygous or homozygous p.G137S A-allele carriers (Figure S4).

**Table 1.** Analyses of association between the p.G137S variant and circulating lipids and cardiovascular health markers.

| Trait | n | $\beta_{SD}(SE)$ | $\beta$ | P |
| --- | --- | --- | --- | --- |
| **Lipid profile** |  |  |  |  |
| Fs LDL cholesterol (mmol/L) | 3938 | 0.67 (0.031) | 0.75 | $5.5 \cdot 10^{-87}$ |
| Fs LDL cholesterol (mmol/L)* | 4077 | 0.67 (0.031) | 0.79 | $1.7 \cdot 10^{-90}$ |
| Fs total cholesterol (mmol/L) | 4496 | 0.56 (0.029) | 0.69 | $1.8 \cdot 10^{-72}$ |
| Fs total cholesterol (mmol/L)* | 4636 | 0.56 (0.029) | 0.71 | $2.5 \cdot 10^{-75}$ |
| Fs HDL cholesterol (mmol/L) | 4631 | -0.11 (0.030) | -0.058 | 0.00041 |
| Fs triglyceride (mmol/L) | 4105 | 0.043 (0.033) | 0.025 | 0.19 |
| Apolipoprotein B (g/L) | 1232 | 0.51 (0.057) | 0.12 | $1.7 \cdot 10^{-17}$ |
| Apolipoprotein A1 (g/L) | 1232 | -0.13 (0.058) | -0.034 | 0.032 |
| Fs VLDL cholesterol (mmol/L) | 2086 | -0.043 (0.045) | -0.0091 | 0.34 |
| Remnant cholesterol (mmol/L) | 3938 | 0.034 (0.034) | 0.0055 | 0.31 |
| Cholesterol-lowering drugs^ | 2870 | 1.54 OR (0.18) | - | 0.016 |
| **Cardiovascular profile** |  |  |  |  |
| Pulse rate (beats per minute) | 1524 | 0.0023 (0.050) | 0.12 | 0.96 |
| Systolic blood pressure (mmHg) | 4133 | -0.014 (0.030) | -0.12 | 0.65 |
| Diastolic blood pressure (mmHg) | 4133 | -0.056 (0.031) | -0.61 | 0.072 |
| Albumin-to-creatinine ratio in urine (mg/L) | 2560 | 0.076 (0.037) | 7.9 | 0.042 |
| Carotid intima-media thickness (mm) | 1063 | 0.080 (0.039) | 0.013 | 0.042 |

\* = including individuals taking cholesterol-lowering drugs.
^ = binary trait.
Each analysis was both run with the phenotype quantile transformed to a standard normal distribution for each sex ( $\beta_{SD}$ ) and without any transformation ( $\beta$ ). The P-values are from the transformed analyses. $\beta$ , effect size; SD, standard deviation; SE, standard error.

**Figure 1.**
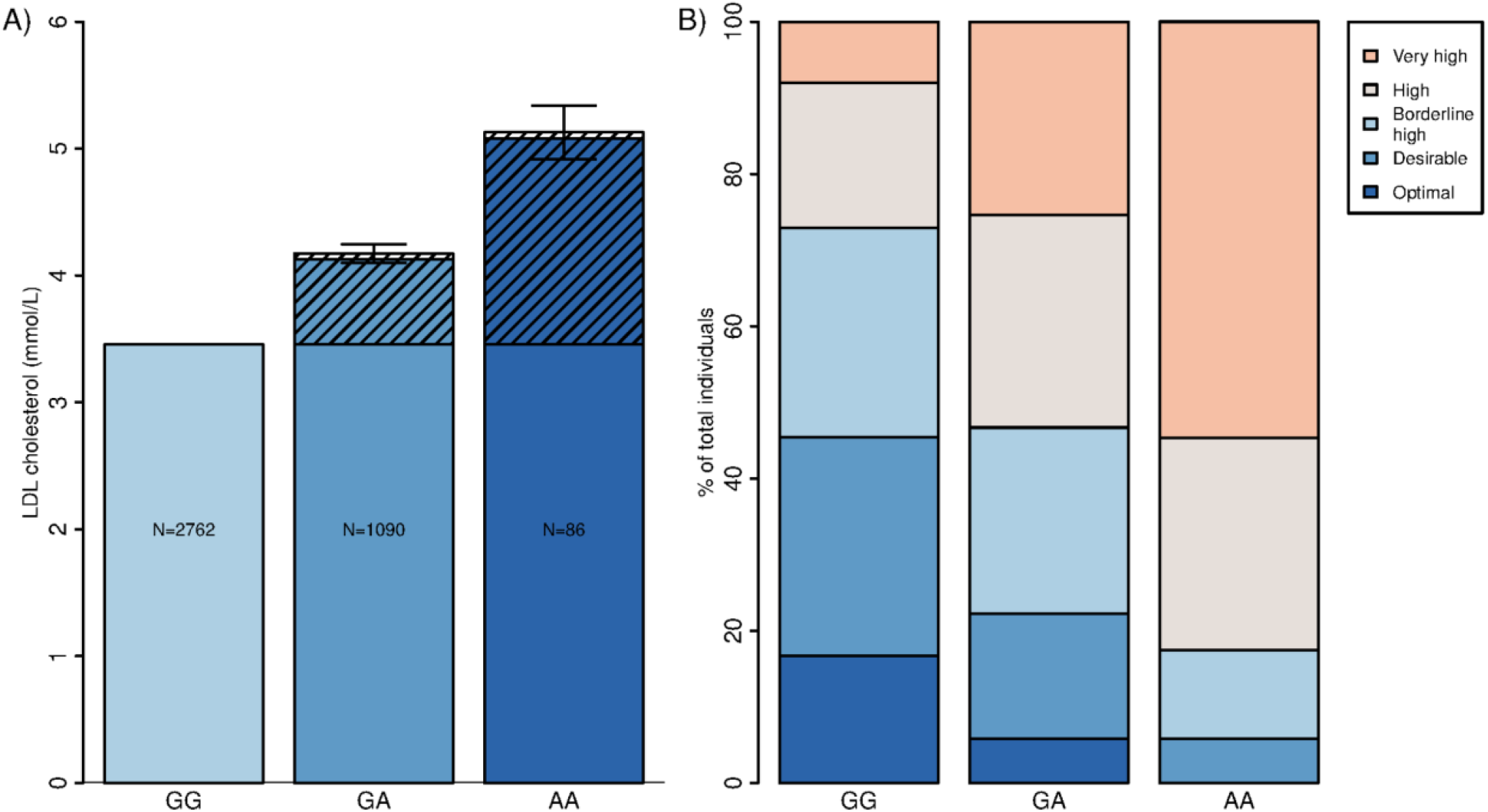
Effect of p.G137S variant on circulating lipid levels. **A)** Mean levels of LDL cholesterol stratified per genotype, as well as estimated effect sizes from GEMMA according to a full model (hatched area) with 95% confidence intervals. The number of participants in each group is written inside the bars. **B)** For each genotype group the proportion of different LDL cholesterol level categories are shown. The LDL cholesterol level categories are according to US guidelines ^45^ with < 2.6 mmol/L being Optimal, 2.6-3.3 mmol/L being Desirable, 3.4-4.0 mmol/L being Borderline high, 4.1-4.8 mmol/L being High and ≥ 4.9 mmol/L being Very high.

We also validated the association between p.G137S and levels of total cholesterol (β, 0.71 mmol/L, P=1.8· 10^−72^, Table 1) and apolipoprotein B (β, 0.12 g/L, P=1.7· 10^−17^, Table 1). Furthermore, we observed a novel association between p.G137S and lower HDL cholesterol levels (β, -0.058 mmol/L P=0.00041, Table 1), and an increased frequency of the use of cholesterol-lowering drugs among risk A-allele carriers (odds ratio (OR) (95% confidence interval (CI)),1.54 (1.08-2.20), P=0.016, Table 1). With respect to basic clinical markers of cardiovascular health, we observed nominally significantly higher levels of urinary albumin-to-creatinine ratio as well as carotid intima-media thickness (Table 1). We found no associations to phenotypes related to body composition (Table S2). Our tests were adjusted for different ancestries through a mixed model and we observed no indication of inflation of the test statistics based on P-value QQ plots for all genotyped variants in the genome (Figure S5).

### Variance explained by the p.G137S variant

Given the large effect size estimates obtained for some of the lipid traits, we estimated the amount of phenotypic variance explained by the variant for these traits. Based on partial R^2^ from a linear model that included PCs, sex, age, and survey, the p.G137S variant explained 11.8% of the variance for LDL cholesterol. Restricting the analyses to 696 individuals with more than 95% Inuit ancestry, the variance explained increased to 16.8% (Figure S6B). We also compared the impact of the variant to that of the main known clinical risk factors for elevated LDL cholesterol, namely BMI, sex, smoking, age, and waist-hip ratio. We found that the p.G137S variant explained more variance of LDL cholesterol than any of these risk factors both when including all risk factors in the model and when estimating partial R^2^ from a model, which only included a single risk factor and the PCs (Figure 2A). To enable comparisons with other variants, we also estimated the variance explained using a less flexible, but commonly used approach based on summary statistics ^28^. This led to similar, but slightly lower estimates of 10.4% variance explained in all individuals and 14.9% in individuals with more than 95% Inuit ancestry only (Figure S6A). Compared to the p.G137S variant, the variants identified in a recent large European GWAS ^29^ had much lower effect sizes and explained much less of the observed variance in LDL cholesterol levels (Figure 2B and 2C).

**Figure 2.**
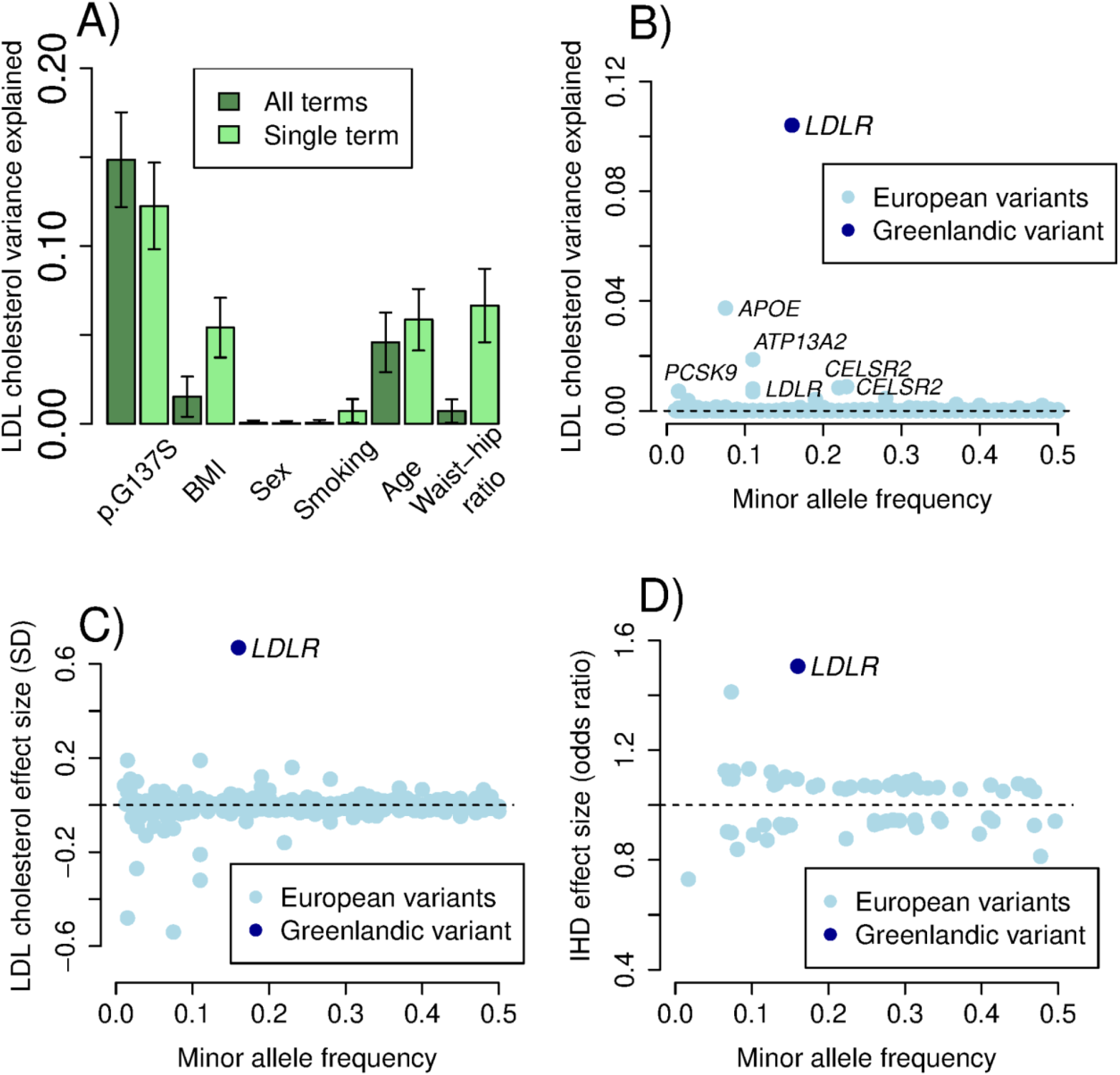
Effect sizes and explained variance of common variants associated with LDL cholesterol and ischemic heart disease in Europeans and the *LDLR* p.G137S variant in Greenlanders. **A)** Comparison of LDL cholesterol variance explained by the p.G137S variant, BMI, sex, smoking, age, and waist-hip ratio, respectively in a model including all terms (All terms) as well as a model including the terms one at a time (Single term), **B**) estimated LDL cholesterol variance (using quantile transformed effect sizes) explained by *LDLR* p.G137S in Greenlanders, and the genetic variants identified to be associated with LDL cholesterol levels in Europeans ^29^ (with gene names), **C**) LDL cholesterol effect sizes for *LDLR* p.G137S in Greenlanders, and the genetic variants identified to be associated with LDL cholesterol levels in Europeans ^29^, and **D)** the odds ratio for ischemic heart disease for *LDLR* p.G137S variant in Greenlanders, and genetic variants identified to be associated with increased risk of ischemic heart disease in Europeans ^30^. Genetic variants with a minor allele frequency < 0.01, have been omitted from the plot.

### Analyses of NMR-based lipoprotein profile and metabolic phenotypes

To explore the effect of the p.G137S variant further, we tested for associations with NMR based measurements of lipoprotein particles and metabolic markers in up to 1520 Greenlanders from our separate Greenlandic cohort. We found a similar effect on overall lipid levels as in the main Greenlandic cohort (Figure S7), however, we also found that variant carriers had a significantly smaller diameter of LDL particles (β_SD_(SE), -0.22 (0.05), P=1.7· 10^−6^), and higher concentrations of all three subclasses of LDL particles (Large, 0.64 (0.04), P=2.6· 10^−48^; Medium, 0.63 (0.04), P=1.8· 10^−46^; Small, 0.62 (0.04), P=1.6· 10^−45^), IDL particles (0.62 (0.04), P=4.7· 10^−45^), and the smallest subclasses of VLDL particles (Small, 0.18 (0.05), 1.6· 10^−4^; Very small, 0.49 (0.04), P=6.6· 10^−29^; Figure 3). In these particles, the content of total lipids, phospholipids, cholesterol, cholesterol esters, free cholesterol, and triglycerides, were all significantly higher in p.G137S carriers (Figure S7). In line with the higher levels of LDL cholesterol, the p.G137S carriers also had higher levels of Apolipoprotein B (0.50 (0.04), 2.9· 10^−29^; Figure 3). Moreover, p.G137S was significantly associated with higher concentration of several cholesterol subtypes, and with a pattern of HDL particles, with significantly higher concentrations of HDL3 cholesterol (0.20 (0.05), P=1.3· 10^−5^), but significantly lower concentrations of large and medium HDL particles (Large, -0.18 (0.05), P=1.3· 10^−4^; Medium, -0.28 (0.05), P=5.7· 10^−9^; Figure 3). With respect to phospholipids and glycerides, the p.G137S carriers had significantly higher concentrations of total phosphoglycerides (0.19 (0.05), P=3.0· 10^−5^), phosphatidylcholine and other cholines (0.19 (0.05), P=3.8· 10^−5^), sphingomyelins (0.45 (0.04), P=5.7· 10^−24^), and total cholines (0.26 (0.04), P=4.7· 10^−9^; Figure 3), and affected some classes of fatty acids. We observed no effect on the concentration of amino acids, or markers of glucose metabolism (Figure S8).

**Figure 3.**
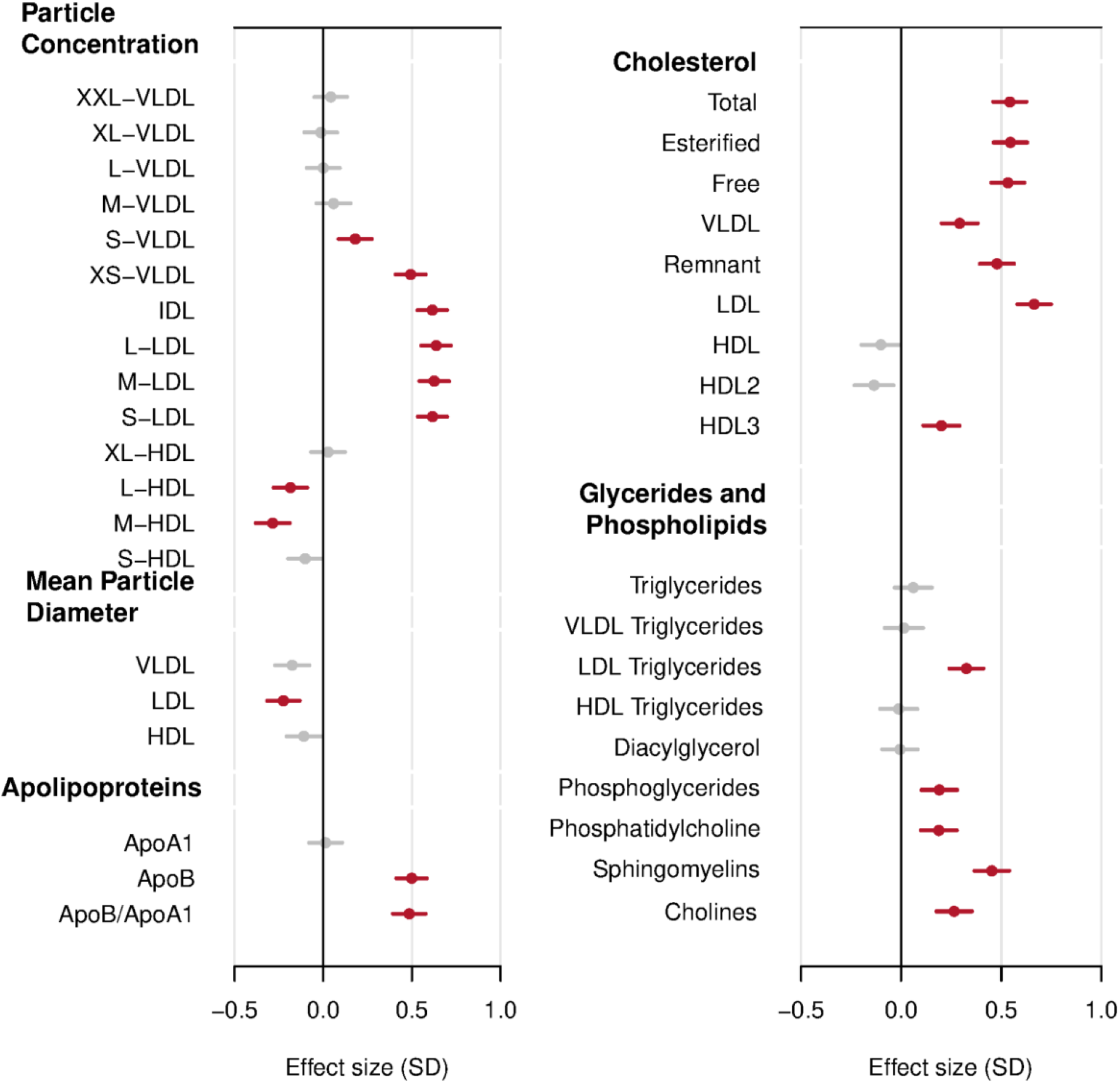
Effect of the p.G137S variant on concentration of lipoprotein particles and cholesterol species. Lipoprotein particle concentration and metabolic measures in Greenlanders with NMR phenotypes. Estimates of the effect size of the p.G137S variant are shown with their 95% confidence interval, furthermore the confidence intervals are coloured red if their P-value is below the bonferroni correction of 3.3· 10^−4^, and grey otherwise. Phenotype values were quantile transformed to a standard normal distribution. XXL, extremely large; XL, very large; L, large; M, medium; S, small; XS, very small; VLDL, very low-density lipoprotein; IDL, intermediate-density lipoprotein; LDL, low-density lipoprotein; HDL, high-density lipoprotein; Apo, apolipoprotein.

### Analysis of cardiovascular disease outcomes

We also analysed if the p.G137S variant had an effect on CVD outcomes in a data set comprising the 4565 individuals, for whom we had both genetic and CVD data.

For the p.G137S carriers we observed a significantly higher risk of ischemic heart disease (HR (95% CI), 1.51 (1.18-1.92), P=0.00096), peripheral artery disease (1.69 (1.01-2.82), P=0.046), and coronary operations (1.78 (1.21-2.62), P=0.0035; Figure 4 and Table S3). For these three types of CVD outcomes, heterozygous and homozygous carriers had lower survival rates as a function of years lived compared to non-carriers (Figure 4, Table S3, Figure S9 and S10). A QQ plot of the test statistics from testing all genotyped variants in the genome showed no indications of inflation of the test statistics in the Cox regression analyses (Figure S11). We observed similar HRs when assessing years since inclusion, instead of years lived (Table S4), and when analysing CVD risk cross-sectionally (Table S5). Remarkably, the observed effect of the p.G137S variant on ischemic heart disease, when doing a cross-sectional analysis in Greenlanders was much greater than the effect of any common variant identified in Europeans ^30^ (Figure 2D). Moreover, these associations remained when adjusting for BMI, smoking, and blood pressure (data not shown). On the contrary, we observed no association with cerebrovascular disease, or a combined measure of the different types of CVD events (Figure 4 & Table S3).

**Figure 4.**
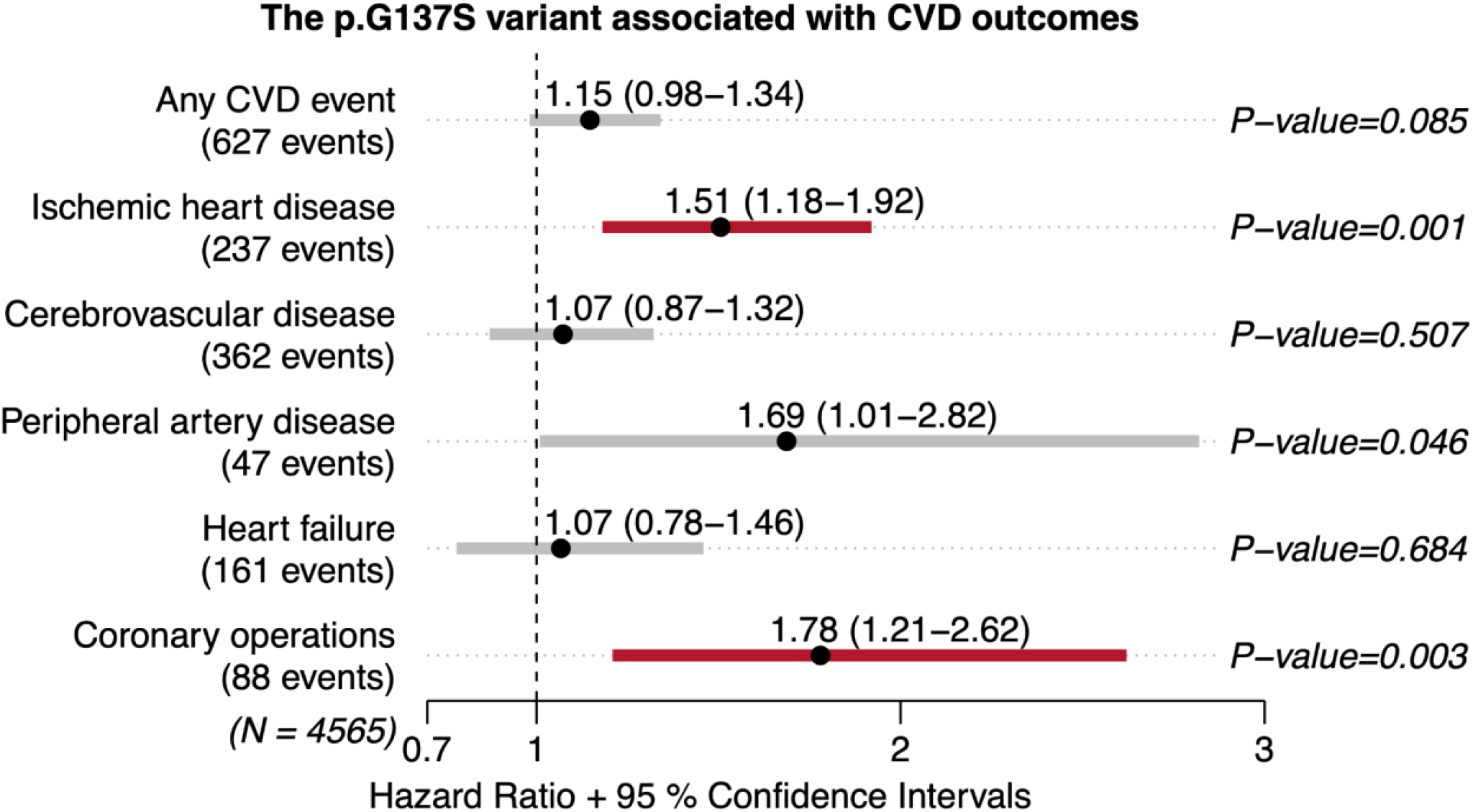
Estimated hazard ratios for the p.G137S variant on CVD. Hazard ratios are shown with their 95% confidence interval and P-values (P) estimated with Cox regression testing for an association between the p.G137S variant and risk of different types of CVD events. The red colour denotes those where the P-value is below the bonferroni correction of 0.00833.

### Investigating through which phenotype the effect on CVD is mediated

Since the p.G137S variant is strongly associated with higher levels of LDL cholesterol, total cholesterol, apolipoprotein B, and lower levels of HDL cholesterol, the effect on CVD risk could be mediated by one of these phenotypes. To investigate if this was the case, we repeated the survival analyses for each of the relevant CVD outcomes with these four lipid phenotypes as covariates one at a time. For ischemic heart disease and peripheral artery disease these analyses showed that the association of the p.G137S variant was still statistically significant after adjusting for HDL cholesterol, and total cholesterol, respectively. However, it was not statistically significant when adjusting for LDL cholesterol for ischemic heart disease and when adjusting for apolipoprotein B for peripheral artery disease (Table S6).

## Discussion

In this study, we investigated the impact of the common Arctic specific *LDLR* p.G137S missense variant on specific lipids as well as metabolic and cardiovascular health in the Greenlandic population.

First, we replicated the previously reported associations with elevated circulating concentrations of LDL cholesterol, total cholesterol, and apolipoprotein B. Notably, our results showed that the p.G137S variant had an even larger impact on the lipid profile of Greenlanders than previously reported ^18^. For LDL cholesterol, we estimated an effect size of 0.75 mmol/L per p.G137S allele, whereas the previously reported estimate was 0.54 mmol/L ^18^. Furthermore, we found an association with HDL cholesterol not previously reported. A likely explanation for these differences is that the previous analyses were performed using a multivariate linear model adjusting only for age, sex, BMI, and geographic location, whereas we used a linear mixed model accounting additionally for population structure and relatedness. In the admixed Greenlandic population this model enabled a more accurate estimation of effect sizes.

Interestingly, the effect size for LDL cholesterol was much larger than the effect sizes reported for any common variant identified in Europeans ^29^. This, combined with the fact that the variant is common and carried by 29.5% of the Greenlandic population, means that the variant has a large impact on this population. This large impact on the population level, was particularly evident by the fact that 25.4% of the heterozygous and 54.7% of the homozygous carriers had LDL cholesterol levels above the diagnostic level for FH and by the fact that the explained variance for LDL cholesterol in Greenlanders was estimated to be around 12%, and more than 16% when only looking at unadmixed Inuit. This estimated amount of variance explained by the *LDLR* p.G137S variant is more than that of any identified common variant in Europeans, and also more than that of clinical risk factors like BMI, sex, smoking, age, or waist-hip ratio explain. For comparison, estimates of the amount of phenotypic variance explained by genetics in large scale European studies using polygenic risk scores range from 10% ^13^ to 19.9% ^31^. Hence, the p.G137S variant alone has an impact in the Greenlandic population comparable to an entire polygenic risk score based on a large study in Europeans.

Thus, the *LDLR* p.G137S variant is an example of a genetic variant with the unusual combination of having a large effect size and being common. This pattern is similar to the common variants with large effects on lipid levels, BMI, or risk of type 2 diabetes in isolated populations of Greeks, Greenlanders, Samoans, or Pima Indians, respectively ^32–37^. Importantly, the *LDLR* variant has an additive effect, and thereby potentially a bigger impact on the general population health as compared to variants with a recessive effect, like the diabetes-causing *TBC1D4* variant previously identified in Greenlanders ^35^.

When we looked further into the effects of the p.G137S variant on the lipid-related measures, from the NMR data, we observed higher concentrations of atherosclerotic lipoproteins of LDL, IDL, and VLDL subclasses, as well as apolipoprotein B in p.G137S carriers, which is in line with reduced function of the LDL receptor. We also observed higher concentrations of HDL3 cholesterol, but lower concentrations of large and medium HDL particles, which might indicate reduced flux of cholesterol through the reverse cholesterol transport system, and thereby reduced hepatic cholesterol uptake. The observed lipoprotein profile in the carriers of the p.G137S carriers was very similar to the profile reported for children with FH with different causative mutations, however, with larger effect sizes in the children ^38^. This difference could be due to the functional effect of the p.G137S variant, approximately reducing the ligand binding to the LDL receptor by 60%, as compared to the FH mutations, which in general have greater functional impact^18,39^.

Regarding the cardiovascular disease impact of the p.G137S variant, we found a significantly increased risk of ischaemic heart disease, coronary operations, and peripheral artery disease, and observed similar effect estimates when limiting the analyses to events after inclusion or when analysing CVD risk cross-sectionally. Importantly, these findings are in line with previous results for patients with FH, who are known to have a higher risk of CVD, especially ischemic heart disease ^9^ but risk of cerebrovascular disease on level with the general population ^13,40^. Generally, our findings were consistent with a recent Mendelian randomisation study where elevated LDL cholesterol levels were shown to be causally linked to increased risk of ischemic heart disease but not ischemic stroke ^41^.

Apolipoprotein B has been found in a recent large Mendelian randomization study to be underlying the causal relationship of circulating blood lipids with ischemic heart disease, in a model of ischemic heart disease conditioned on apolipoprotein B and respectively LDL cholesterol and triglycerides or apolipoprotein A1 and HDL cholesterol ^42^. For the p.G137S variant, our results failed to indicate that the level of apolipoprotein B was a better predictor of increased risk of ischemic heart disease, as the HRs were reduced similarly when adjusting for LDL cholesterol. Interestingly, the NMR analyses revealed alternative lipid species, which could be contributing to the increased risk of CVD outcomes in p.G137S carriers. Hence, in these individuals, we observed higher concentrations of both phosphatidylcholines and sphingomyelins, the latter class including ceramides. Specific subtypes of these lipid species have previously been linked to increased CVD risk, and these associations seem to be independent of other atherosclerotic risk factors, including age, BMI, smoking, triglycerides, as well as LDL and total cholesterol ^43^. Extended lipidomics profiling assessing specific phosphatidylcholines and ceramides further elucidating these possibly causal relationships would be of great interest.

Only around 10% of the homozygous p.G137S carriers were prescribed cholesterol-lowering drugs, even though most of them had LDL cholesterol levels above 4.9 mmol/L, which is the level considered diagnostic for FH ^9^. This suggests that there are many currently untreated individuals who potentially could benefit from treatment. Moreover, we observed that the elevated levels of LDL cholesterol for p.G137S carriers was independent of age, indicating that these individuals would benefit from early intervention and treatment, similar to the recommendations for treatment of FH ^9^. It has been shown for FH individuals that early statin therapy can help mitigate their risk of developing IHD so that it is almost similar to individuals without FH ^15,44^. Our findings coupled with the fact that the p.G137S variant is common among Greenlanders, indicate that a screening program for the p.G137S variant could be highly useful for early identification of individuals at increased CVD risk, hence, potentially improving preventive care and public health.

In summary, the common p.G137S variant had a major effect on the circulating lipid profile of Greenlanders, where carriers were characterised by an atherogenic lipid profile including elevated levels of LDL cholesterol, total cholesterol, and apolipoprotein B as well as an increased risk of CVD. Furthermore, the large effect size for LDL cholesterol combined with the high frequency of the variant, mean that the variant has a large population level impact. The variant is therefore a simple marker that could be used as an early indicator of future risk of increased lipid levels and CVD. Early detection of individuals likely benefiting from intervention with cholesterol-lowering drugs, could potentially lead to improved CVD prevention in Arctic populations.

## Data Availability

The Greenlandic Metabochip-genotype data are deposited in the European Genome-phenome Archive
(https://www.ebi.ac.uk/ega/home) under the accessions EGAS00001002641

## Acknowledgments

We gratefully acknowledge the participants in the Greenlandic health surveys.

