## Supplementary material for "An LDLR missense variant poses high risk of familial hypercholesterolemia in 30% of Greenlanders and offers potential for early cardiovascular disease intervention"

**Figure S1.** Boxplot of gene expression of LDLR and the p.G137S variant
**Figure S2.** Association between LDL cholesterol levels and the p.G137S variant by age group
**Figure S3.** Association between LDL cholesterol levels and the p.G137S variant by admixture group
**Figure S4.** Boxplot of LDL cholesterol levels (mmol/L) for each of the three genotype based groups
**Figure S5.** QQ-plots for association analyses of the four significantly associated metabolic traits using a linear mixed model
**Figure S6.** Estimates of the amount of variance in LDL cholesterol (LDLC), total cholesterol (CHOL), HDL cholesterol (HDLC) and apolipoprotein B (apoB) explained by the p.G137S variant
**Figure S7.** Association between lipid NMR phenotypes and the p.G137S variant
**Figure S8.** Association between metabolic NMR phenotypes and the p.G137S variant
**Figure S9.** Kaplan Meier plots for four types of CVD events
**Figure S10.** Survival plots of the results from the multivariate Cox regression from Figure 4
**Figure S11.** QQ-plot for the Cox regression analyses of the CVD events

**Table S1.** Frequency of the p.G137S variant in populations from across the world
**Table S2.** Association between the p.G137S variant and measures of body composition
**Table S3.** Association between the p.G137S variant and cardiovascular disease outcomes
**Table S4.** Association between the p.G137S variant and cardiovascular disease outcomes, only using events after the inclusion in the study
**Table S5.** Association between the p.G137S variant and cardiovascular disease outcomes, using a logistic mixed model (GMMAT)
**Table S6.** Hazard ratio for selected CVD events with and without adjustment for circulating lipid concentrations
**Table S7.** Classification codes for cardiovascular disease events
**Table S8.** Classification codes for operational procedures related to cardiovascular health

### Supplementary Methods

#### Ethical statement

This study was approved by the Commission for Scientific Research in Greenland (project 2011–13, ref. no. 2011–056978; project 2013–13, ref.no. 2013–090702, and project 2013-17), and was conducted in accordance with the ethical standards of the Declaration of Helsinki, second revision. All participants gave informed consent.

#### Metabolic phenotypes

Serum cholesterol, HDL cholesterol, and triglyceride concentrations were measured using enzymatic calorimetric techniques (Hitachi 917, Roche Molecular Biochemicals, Indianapolis, IN, USA). VLDL and LDL cholesterol concentrations were calculated according to Friedewald's formula. Remnant cholesterol was calculated as total cholesterol subtracting LDL cholesterol and HDL cholesterol. Apolipoprotein A and apolipoprotein B levels were measured using turbidimetric kits using a Cobas Mira S autoanalyzer from Roche Diagnostics (Laval, Quebec, Canada). For LDL cholesterol and serum cholesterol, the concentrations were measured in all individuals, information on cholesterol lowering medicine was obtained by interview and used for excluding those on cholesterol lowering medicine. Participants were standing upright while waist and hip circumference were measured. Waist circumference was measured midway between the rib cage and the iliac crest, and hip circumference was measured at its maximum. Height and weight were measured with the participants wearing underwear and socks and BMI was calculated as weight (kg) divided by height squared ($m^2$).

#### CVD data

Fatal and non-fatal CVD diagnoses and procedures included ischemic heart disease, cerebrovascular diseases, heart failure, and peripheral artery disease. Diagnoses and procedures were obtained from The Greenlandic National Patient register, the Electronic Patient Record in Greenland (COSMIC), the Danish National Patient Register, and the Causes of Death Register from both countries, using the International Classification of Diseases (ICD-) 8, ICD10, and International Classification of Primary Care 2 (ICPC-2) codes, The Classification of Operations and Treatments, and The Nordic Medico-Statistical Committee (NOMESCO) Classification of Surgical Procedures for procedures (Table S7 & S8). The Central Person Register provided data on emigration and death. Validity of cardiovascular diagnoses in both registries, and the validity of cardiac procedures in The Danish National Patient register, have previously been found to be high [1–3]. CVD data was extracted for the main cohort. This provided a data set of 4565 individuals with CVD data and genotype data.

#### NMR data

Serum metabolites were characterized with a high-throughput NMR metabolomics platform [4,5]. This method provides quantitative molecular measures on selected lipoprotein subclasses, their lipid composition and concentrations, apolipoprotein A-I and B, measures of cholesterol and triglyceride, some fatty acids, albumin, as well as on a range of low-molecular-weight metabolites, including glycolysis related molecules, amino acids, and ketone bodies.

#### Estimation of variance explained by p.G137S

To estimate how much of the phenotypic variance is explained by the p.G137S variant for LDL cholesterol, total cholesterol, and HDL cholesterol, we used two approaches. First, we used partial $R^2$ from a linear model with other predictors as well as the top 10 genetic principal components (PCs), age, sex and survey. This approach estimated the variance explained by one predictor after removing the effect of the other predictors. Second, we used the summary statistic based approach of using the formula from [6] as this can be used with estimates from a linear mixed model (LMM), and therefore makes comparisons with other variants from previous studies possible. Note that when using this approach the variance explained can only be estimated for the genotype. The confidence intervals on the estimates for partial $R^2$ is based on 1000 bootstraps and for the LMM we used the standard error on the estimated effect size of the genotype for the variance explained standard error.

**RNA expression data**

Whole transcriptome RNA was extracted from peripheral blood from a subset of individuals (n = 499) from the IHIT and B99 survey. The RNA was then sequenced using the BGI500 sequencing technology (100bp paired-end sequencing). A total of 17–49 (median: 21) million read pairs passed the quality filters and were used for expression quantification. We mapped the reads to GRCh38 (Ensemble v.94 annotation) using STAR [7]. Gene level quantification was obtained using RNA-SeQC [8]. Counts were normalized across samples using TMM [9], followed by quantile normalization per gene. The procedures used for extraction, sequencing and quality filtering have previously been described in detail [10].

**Analysis of RNA data**

We tested for association between gene expression levels for a set of genes neighboring the *LDLR* p.G137S variant by applying a linear mixed model, as implemented in GEMMA [11], where we accounted for genetic relatedness and admixture. Sex, age, and top 10 PCs from a PCA of the normalized expression matrix were included as covariates in the analyses.

**Supplementary Figures**

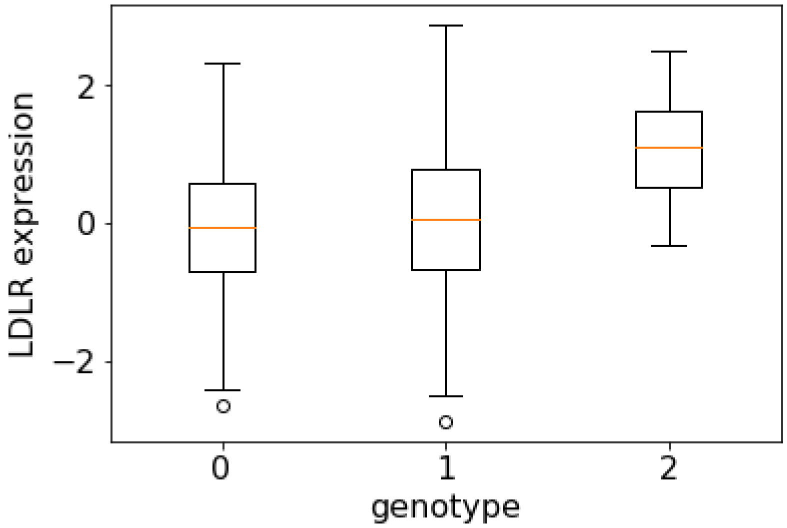


**Figure S1.** A boxplot of gene expression of *LDLR* and the p.G137S variant. Analysis of 494 individuals with RNA data and p.G137S genotype data. A LMM was used to statistically test if the p.G137S variant is an eQTL for *LDLR* the P-value of the test was 0.22.

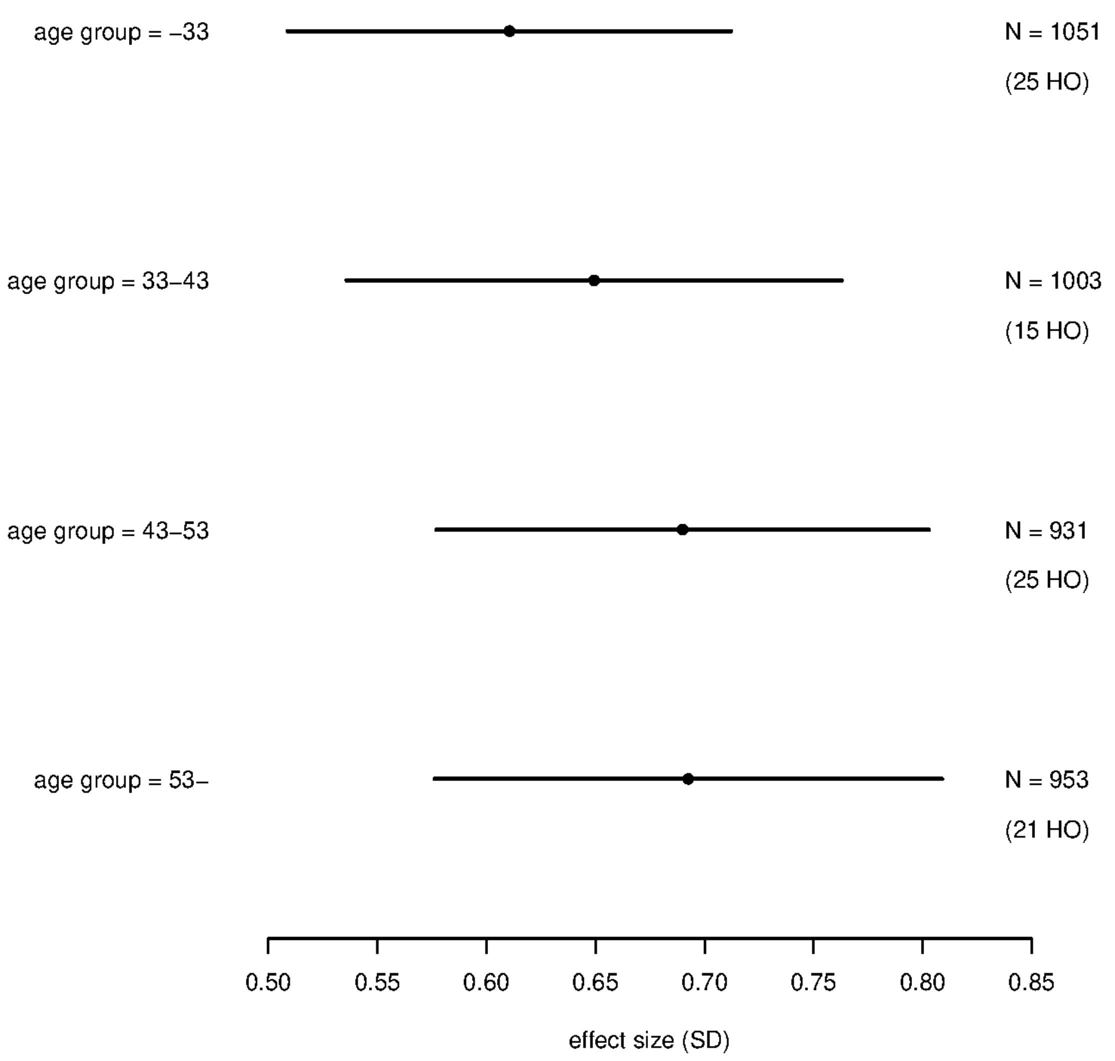


**Figure S2.** Results of testing for an association between LDL cholesterol levels and the p.G137S variant using a linear mixed model dividing the individuals into four approximately equally big groups based on age. The effect size estimate is shown with its 95% confidence interval. We have quantile transformed LDL cholesterol levels to a standard normal distribution for each sex. The number of individuals and homozygous risk carriers for each strata are indicated (N = Number of individuals within each strata, HO = number of homozygous p.G137S variant carriers within each category).

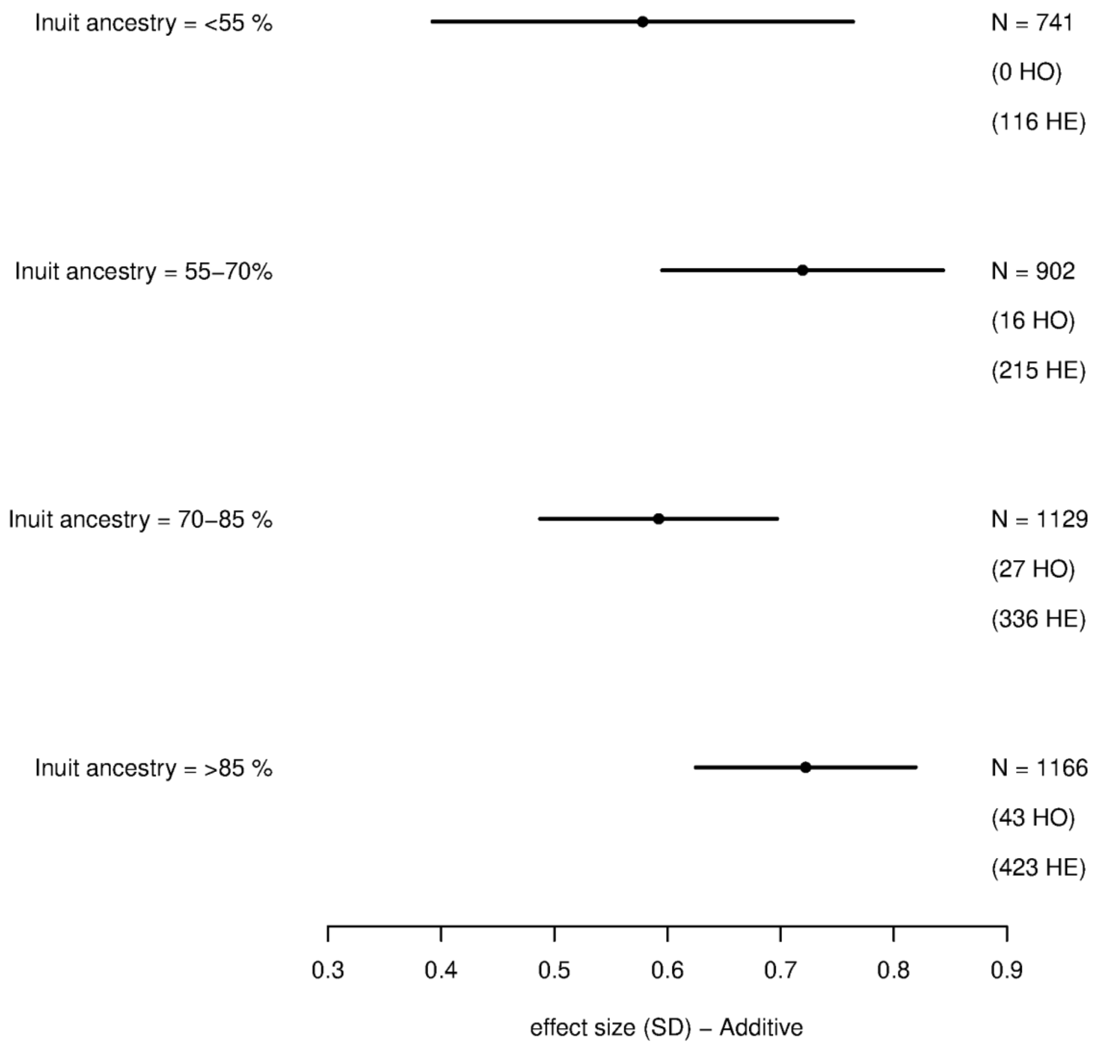


**Figure S3.** Results of testing for an association between LDL cholesterol levels and the p.G137S variant using a linear mixed model dividing the individuals into four groups based on the degree of admixture. The effect size estimate is shown with its 95% confidence interval. We have quantile transformed LDL cholesterol levels to a standard normal distribution for each sex. Effect sizes and 95% confidence intervals are shown for transformed data. The number of individuals and homozygous risk carriers for each strata are indicated (N = Number of individuals within each strata, HE = number of heterozygous p.G137S variant carriers within each category, HO = Same as HE but number of homozygous).

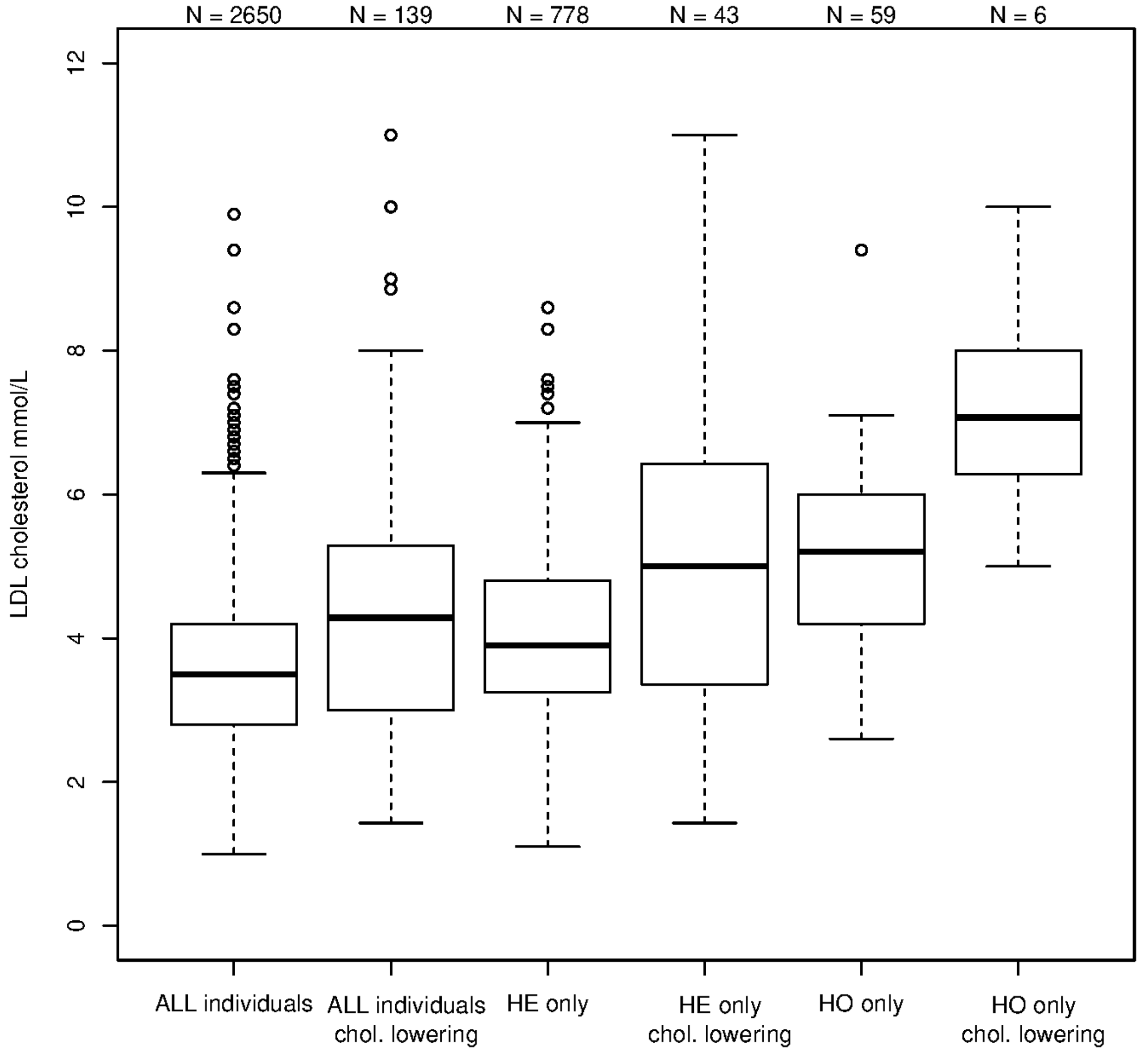


**Figure S4.** Boxplot of LDL cholesterol levels (mmol/L) for different subsets of the data. For each of the three genotype based groups: all individuals (ALL), the heterozygous carriers of p.G137S (HE) and the homozygous carriers of p.G137S (HO) we show the LDL cholesterol levels for those in the group that are not on cholesterol-lowering drugs and those that are on cholesterol-lowering drugs. A linear mixed model was run to test if there is statistically significant difference in LDL cholesterol levels between individuals who are and who are not on cholesterol-lowering drugs, within each group (ALL, HE & HO). ($\beta_{ALL}$(SE), 0.47 (0.09) mmol/L, $P_{ALL}$= $2.37 \cdot 10^{-7}$, $\beta_{HE}$(SE), 0.61 (0.17) mmol/L, $P_{HE} = 3.74 \cdot 10^{-4}$ , & $\beta_{HO}$(SE), 1.45 (0.52) mmol/L, $P_{HO}$ = 0.010). These analyses were performed in the same manner as the main analyses described in the Methods and Materials section, but where the genotype has been replaced with whether an individual is on cholesterol-lowering drugs.

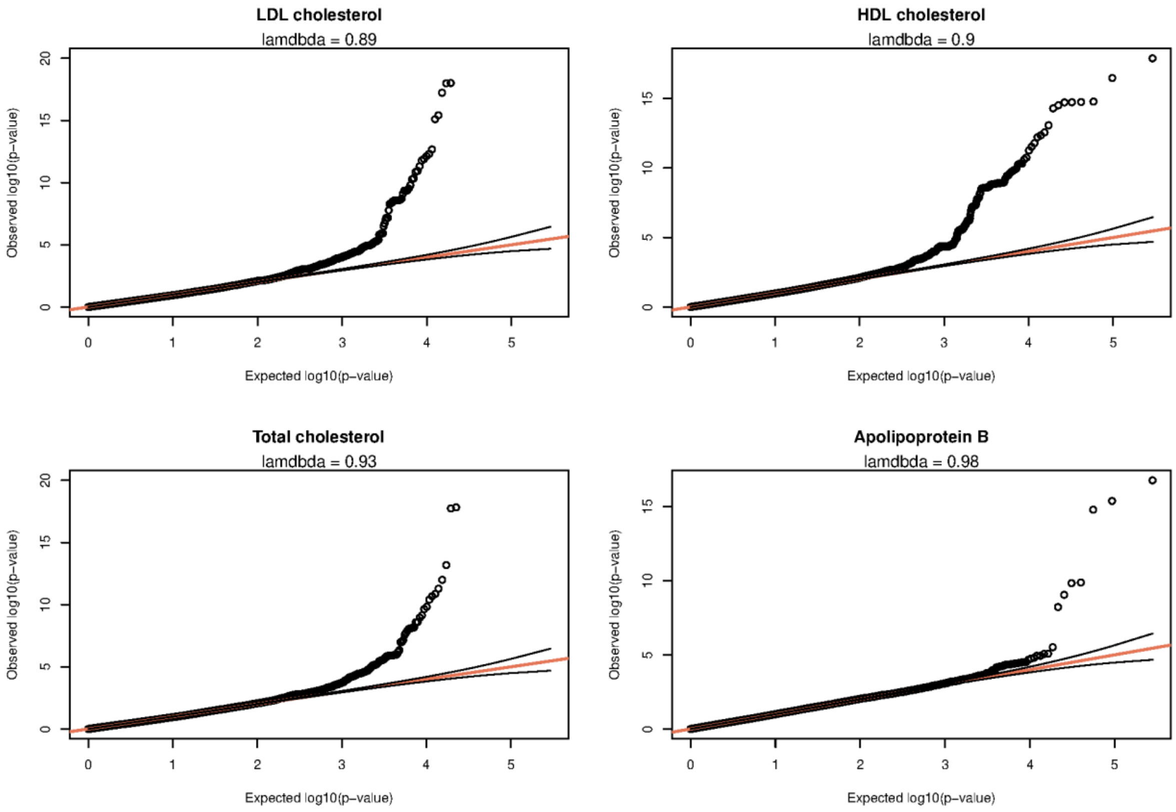


**Figure S5.** QQ-plots for association analyses of the four significantly associated metabolic traits using a linear mixed model. The phenotypes were quantile transformed to a standard normal distribution for each sex and carried out as described Methods and Materials section.

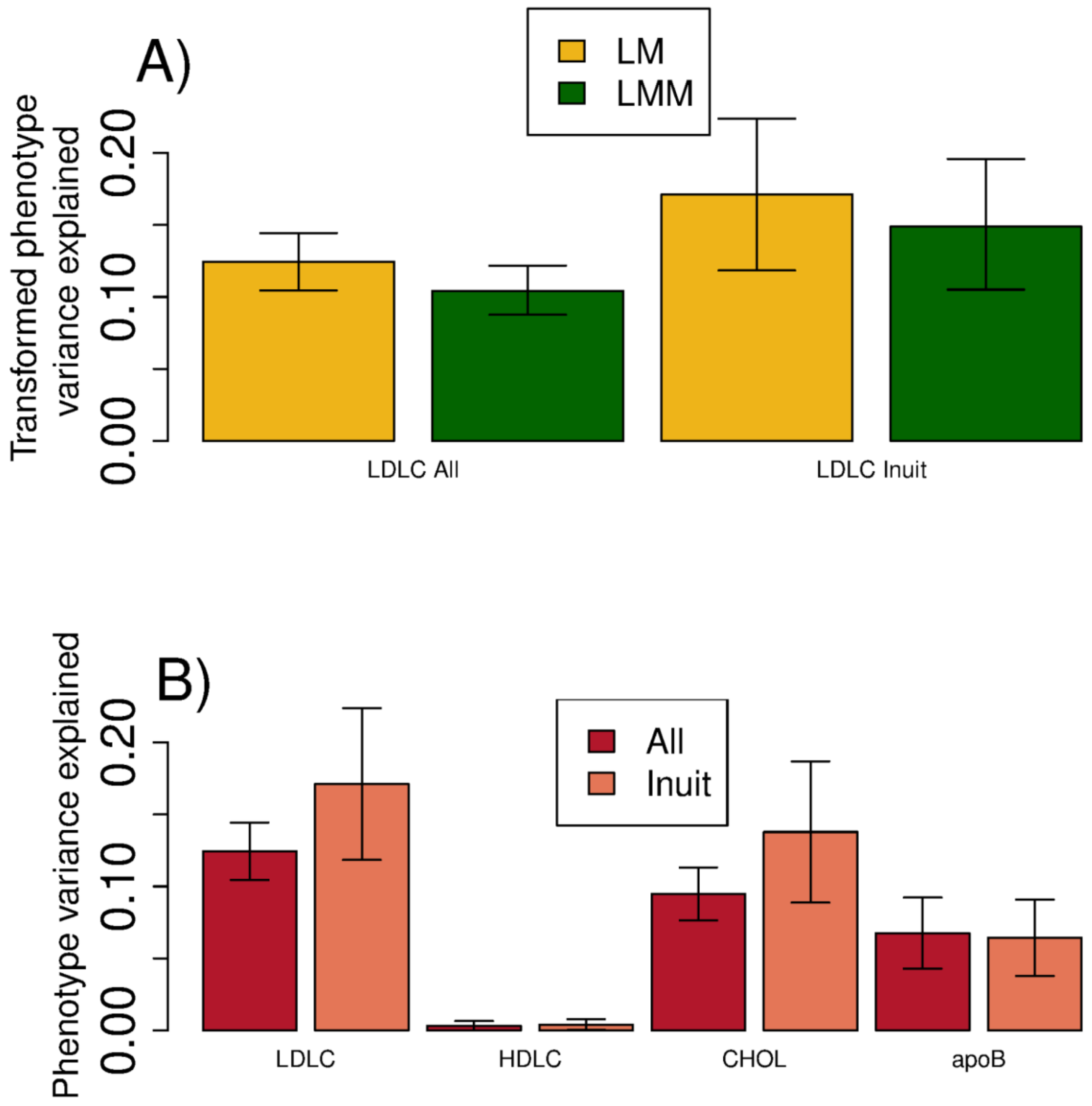


**Figure S6. A)** Estimates of the amount of variance in LDL cholesterol (LDLC) explained by the p.G137S variant obtained using two different approaches. In the first approach the estimates were obtained using a linear model (LM) adjusted for age, sex and the top 10 genetic principal components, and survey, and then using partial $R^2$. In the second approach, based on summary statistics, GEMMA, a linear mixed model (LMM) was used to obtain estimates, which were then plugged into the formula from Shim *et al.*[6] For both approaches, we obtained estimates both by analysing the full cohort and by only analysing individuals with > 0.95 Inuit ancestry. **B)** Variance explained by the p.G137S variant for LDL cholesterol (LDLC), HDL cholesterol (HDLC), total cholesterol (CHOL) and apolipoprotein B, respectively, in the entire cohort and in individuals with >0.95 Inuit ancestry. These estimates are based on using a linear model like in **A)**.

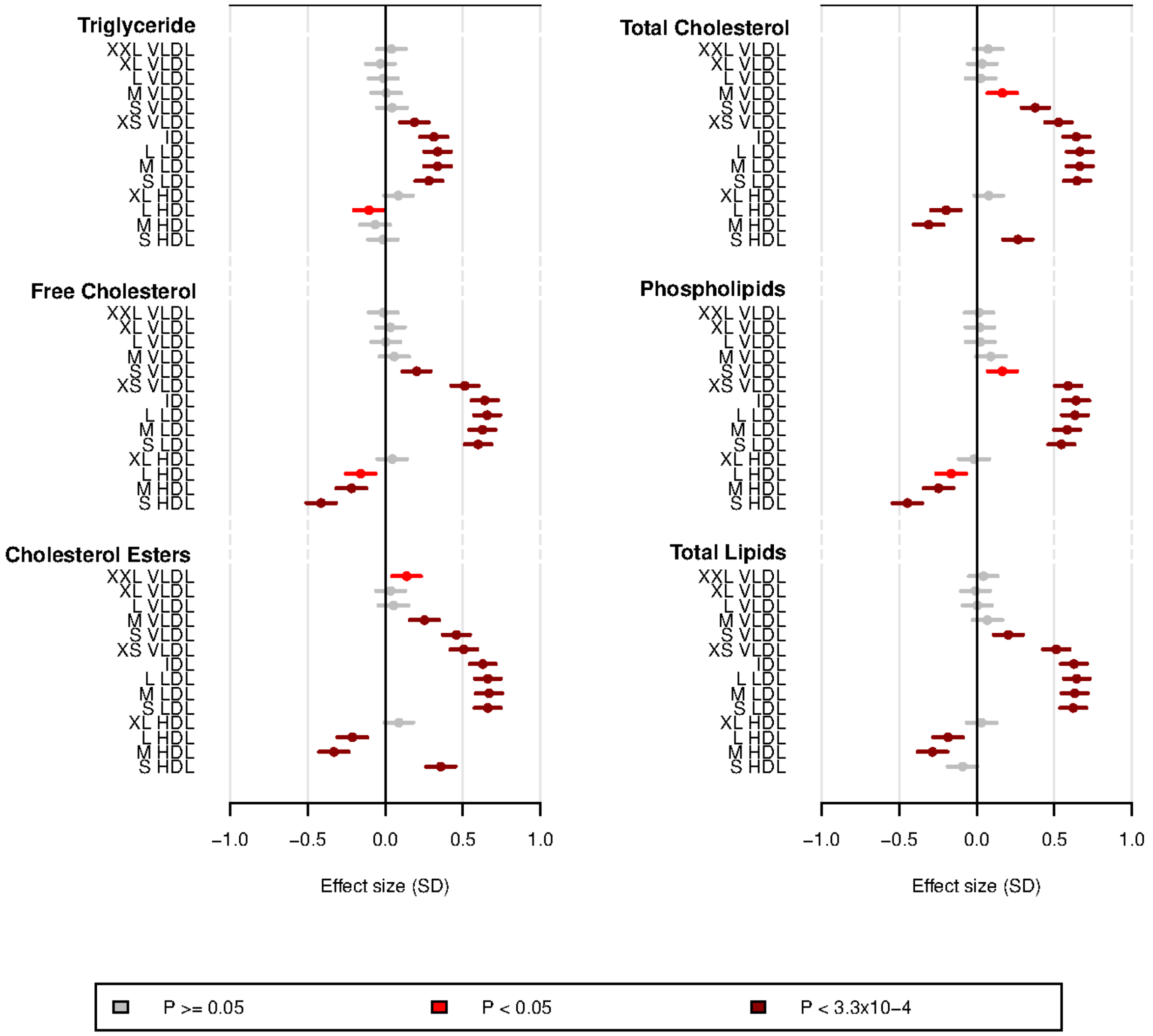


**Figure S7.** Further NMR phenotypes, measuring respectively the triglyceride, free cholesterol, cholesterol esters, total cholesterol, phospholipids and total lipids content of cholesterol particles. Estimates of the effect size of the p.G137S variant are shown with their 95% confidence interval, furthermore the confidence intervals are coloured according to their P-value. Phenotype values were quantile transformed to a standard normal distribution. XXL, extremely large; XL, very large; L, large; M, medium; S, small; XS, very small; VLDL, very low-density lipoprotein; IDL, intermediate-density lipoprotein; LDL, low-density lipoprotein; HDL, high-density lipoprotein.

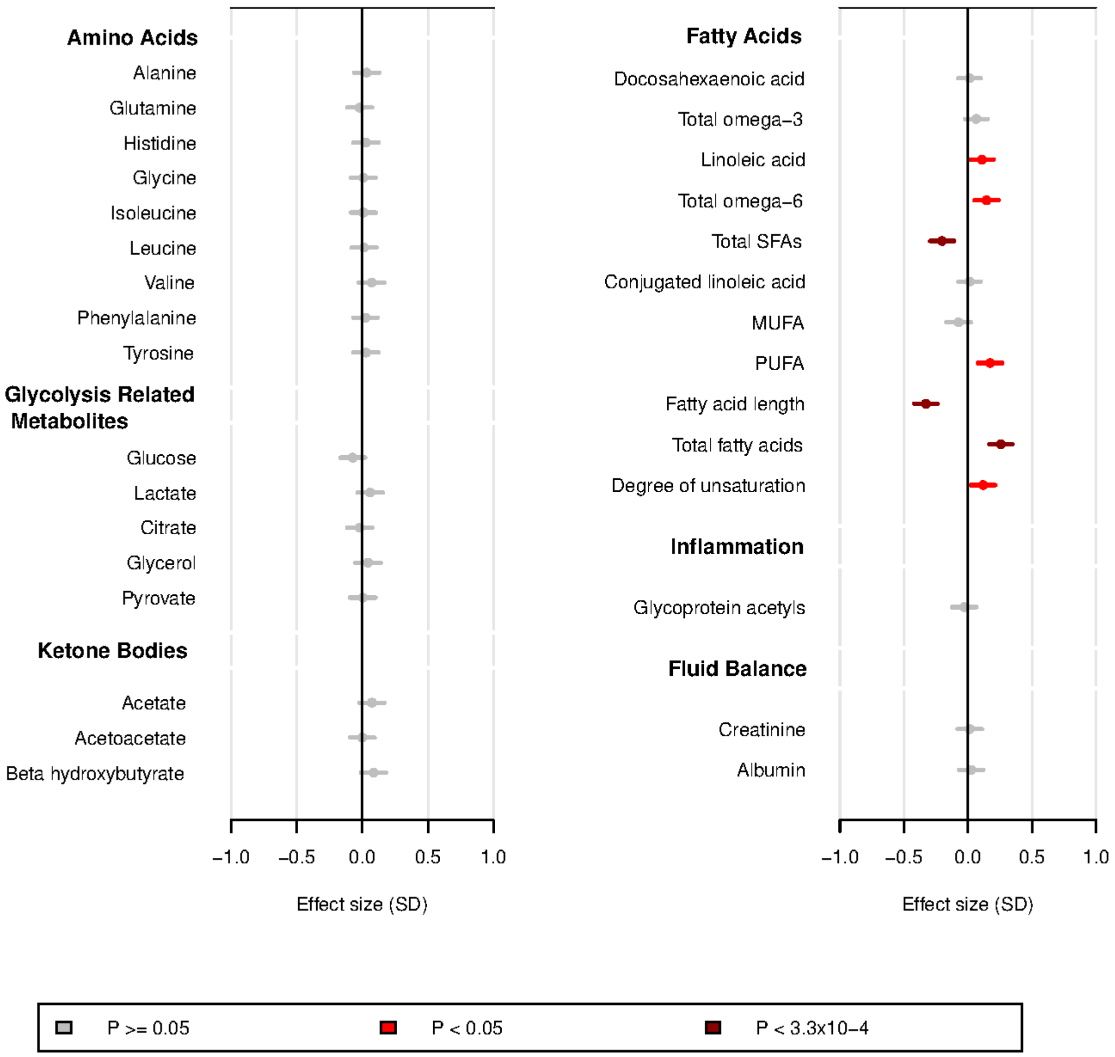


**Figure S8.** Metabolic measures in Greenlanders with NMR phenotypes.
SFA, saturated fatty acids; MUFA, monounsaturated fatty acids; PUFA, polyunsaturated fatty acids. The analyses were done in the same way as for Figure S6.

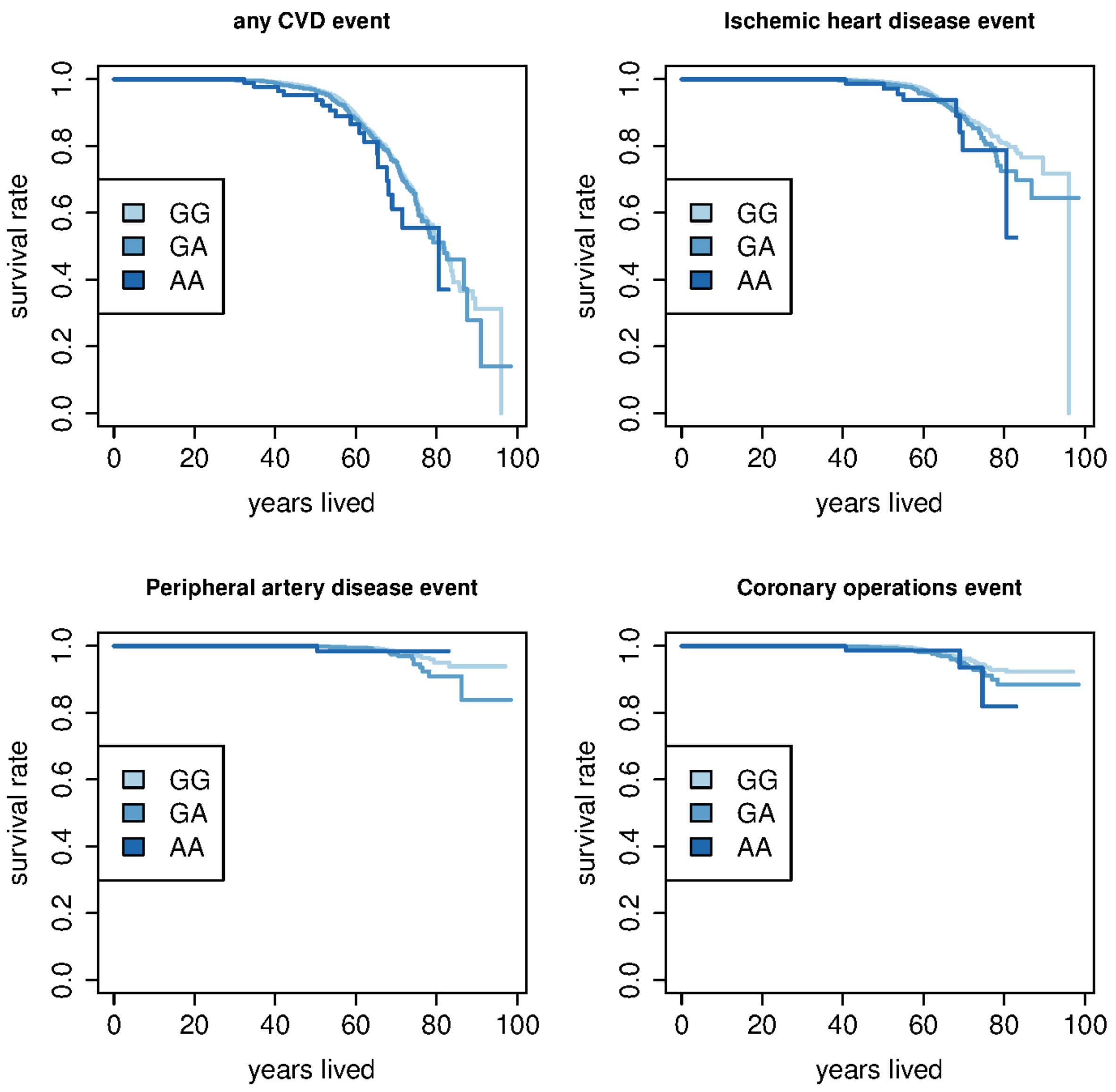


**Figure S9.** Kaplan Meier plots for four types of CVD events. The plots show the survival rate over time for each of the three p.G137S variant genotypes with regards to having had an event of the different types. The types considered are any CVD event, ischemic heart disease, peripheral artery disease and coronary operations.

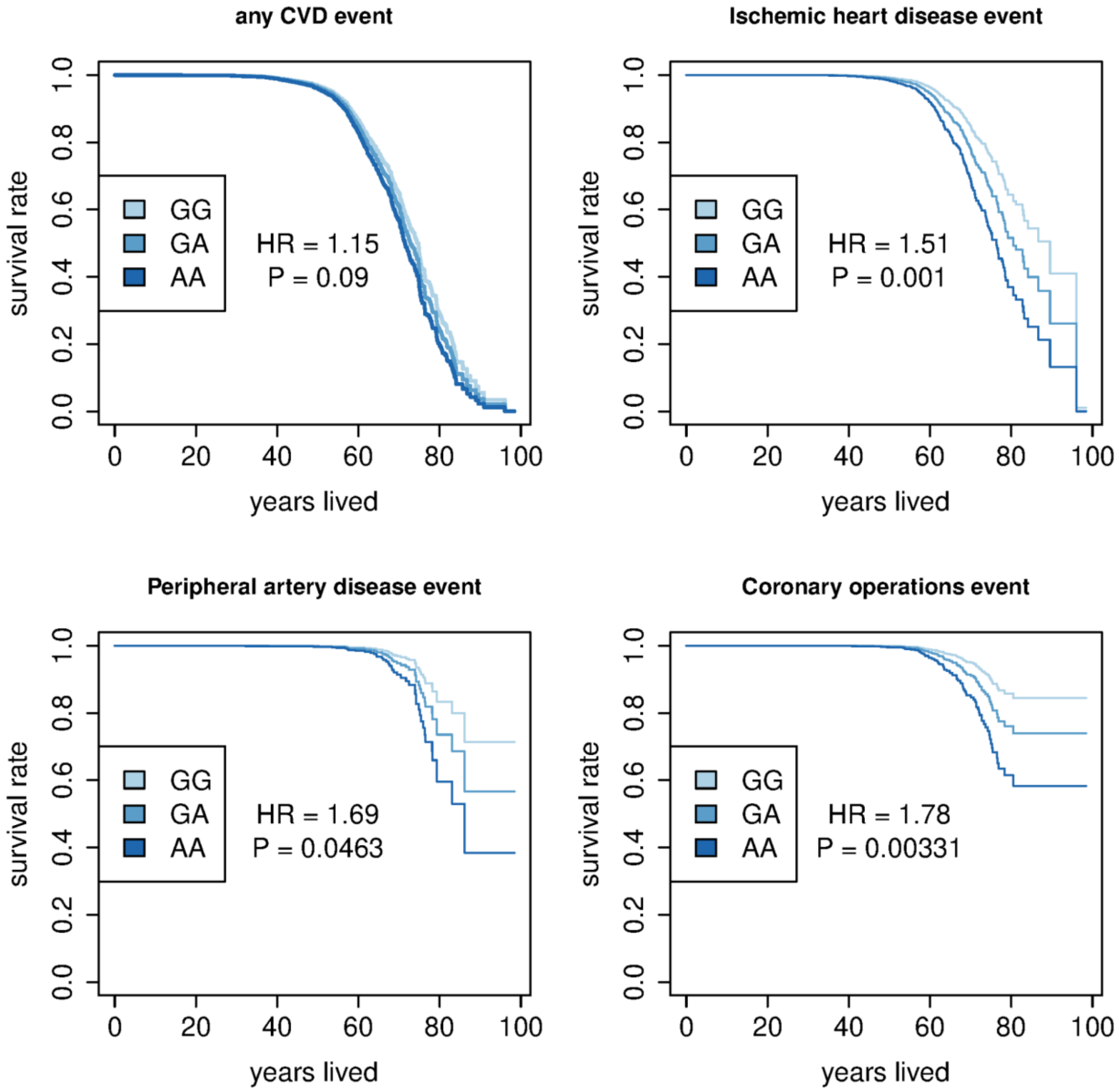


**Figure S10.** Survival plots of the results from the multivariate Cox regression from Figure 4. It shows the predicted survival proportion at any given value of years lived for carriers of the p.G137S carriers with an additive model, where other covariates are fixed to their average value.

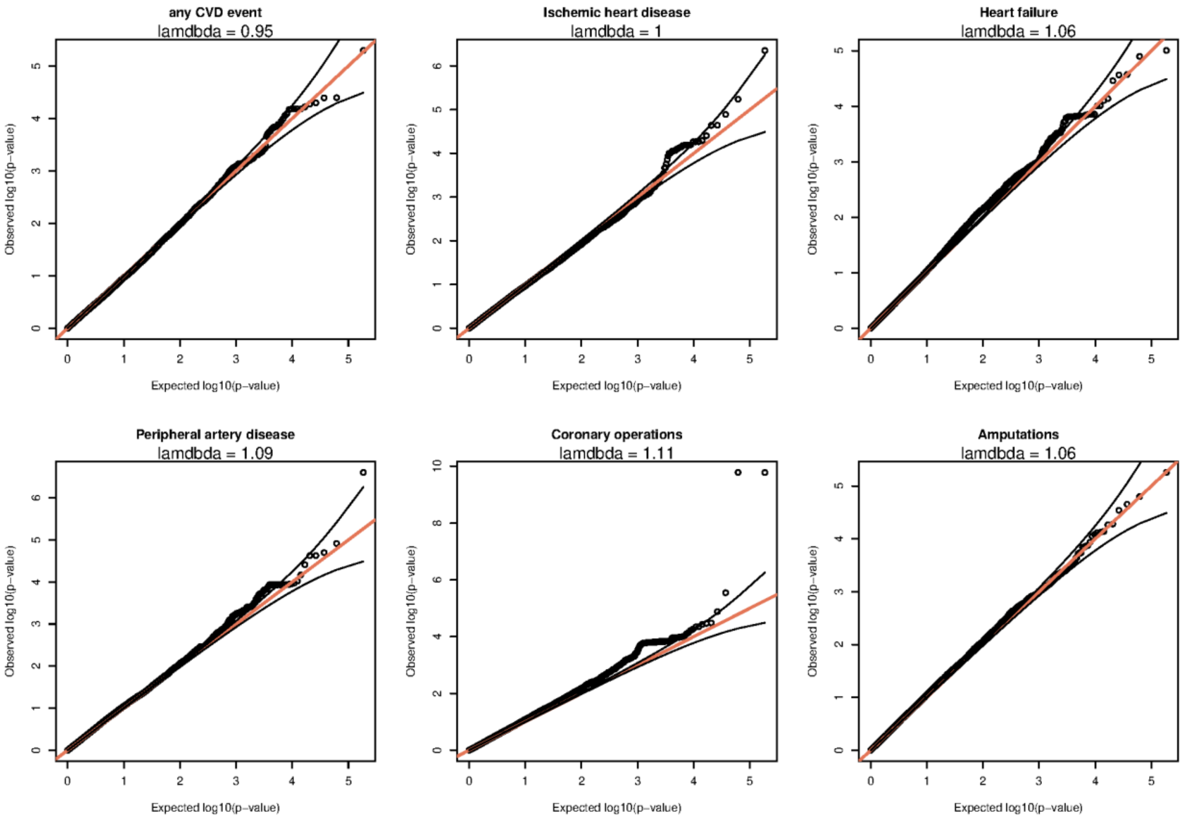


**Figure S11.** QQ-plots for the Cox regression analyses from Figure 4 of CVD events, but for all the variants that passed filters on the MetaboChip.

### Supplementary Tables

#### Table S1. Frequency of the p.G137S variant in populations from across the world

| Region | n | Allele Frequency | Reference |
|---|---|---|---|
| **Americas** | | | |
| Greenlanders | 4653 | 0.158 | This study |
| Ancient Dorset | 16 | 0.00 | Raghavan et al. 2014 |
| Ancient Saqqaq | 2 | 0.00 | Rasmussen et al. 2010 |
| Americas (SGDP) | 27 | 0.00 | SGDP |
| Americas (HGDP) | 51 | 0.00 | HGDP |
| Americas (1000 Genomes) | 347 | 0.00 | 1000 Genomes |
| Latino (gnomAD v3.0) | 6835 | 0.00 | gnomAD v3.0 |
| Latino (gnomAD v2.1) | 34588 | 0.00 | gnomAD v2.1 |
| **Siberia** | | | |
| Central Asia Siberia (SGDP) | 27 | 0.00 | SGDP |
| Central Asia Siberia (HGDP) | 23 | 0.00 | HGDP |
| **Unknown** | | | |
| Other* (gnomAD v2.1†) | 6132 | 0.000082 | gnomAD v2.1 |
| Other* (gnomAD v3.0) | 1077 | 0.00 | gnomAD v3.0 |
| **Rest of the World** | | | |
| Ashkenazi Jew (gnomAD v2.1†) | 10072 | 0.000050 | gnomAD v2.1 |
| South Asian (gnomAD v2.1†) | 30616 | 0.000016 | gnomAD v2.1 |

| | | | |
|---|---|---|---|
| European (non-Finnish) (gnomAD v2.1†) | 113604 | 0.000004 | gnomAD v2.1 |
| Genome Asia 100K | 1739 | 0.00 | 1000 Genomes |
| HGDP | 754 | 0.00 | HGDP |
| SGDP | 246 | 0.00 | SGDP |
| 1000 Genomes | 2157 | 0.00 | 1000 Genomes |

* = This category includes individuals who are in unassigned populations. † = This category includes exome data only. SGDP = Simons Genome Diversity Project, HGDP = Human Genome Diversity Project.

### Table S2. Association between the p.G137S variant and measures of body composition

| Trait | $\beta_{SD}$ (SE) | β | P | n |
|---|---|---|---|---|
| **Body composition** | | | | |
| BMI (kg/m$^2$) | -0.021 (0.031) | -0.12 | 0.506 | 4605 |
| Height (cm) | -0.0098 (0.026) | -0.035 | 0.71 | 4627 |
| Weight (kg) | -0.030 (0.031) | -0.50 | 0.33 | 4610 |
| Waist (cm) | -0.037 (0.030) | -0.49 | 0.22 | 4574 |
| Hip (cm) | -0.0560 (0.073) | -0.54 | 0.073 | 4572 |
| Waist-hip ratio | 0.0025 (0.028) | 0.00058 | 0.93 | 4571 |

All body composition traits tested for an association with the p.G137S variant. Each analysis was both run with the phenotype quantile transformed to a standard normal distribution for each sex ($\beta_{SD}$) and without any transformation (β). The P values are from the transformed analyses.

### Table S3. Association between the p.G137S variant and cardiovascular disease outcomes

| Trait | n | Number of events | HR | 95% CI | P |
|---|---|---|---|---|---|
| Any CVD event | 4565 | 627 | 1.15 | 0.98 - 1.34 | 0.085 |
| Ischemic heart disease | 4565 | 237 | 1.51 | 1.18 - 1.92 | 0.00096 |
| Cerebrovascular disease | 4565 | 362 | 1.07 | 0.87 - 1.32 | 0.51 |
| Peripheral artery disease | 4565 | 47 | 1.69 | 1.01 - 2.82 | 0.046 |

| | | | | | |
|---|---|---|---|---|---|
| Heart failure | 4565 | 161 | 1.07 | 0.78 - 1.46 | 0.68 |
| Coronary operations | 4565 | 88 | 1.78 | 1.21 - 2.62 | 0.0035 |

Hazard ratio (HR), 95% confidence interval (95% CI), and P-values (P) estimated with Cox regression testing for an association between the p.G137S variant and risk of different types of CVD events.

**Table S4. Association between the p.G137S variant and cardiovascular disease outcomes, only using events after the inclusion in the study**

| Trait | n | Number of events | HR | 95% CI | P |
|---|---|---|---|---|---|
| Any CVD event | 4200 | 389 | 1.17 | 0.96 - 1.42 | 0.13 |
| Ischemic heart disease | 4325 | 158 | 1.52 | 1.13 - 2.05 | 0.0062 |
| Cerebrovascular disease | 4277 | 225 | 1.15 | 0.8799 - 1.489 | 0.31 |
| Peripheral artery disease | 4376 | 35 | 1.72 | 0.94 - 3.17 | 0.079 |
| Heart failure | 4351 | 109 | 0.90 | 0.60 - 1.35 | 0.61 |
| Coronary operations | 4366 | 62 | 1.69 | 1.06 - 2.71 | 0.029 |

Hazard ratio (HR), 95% confidence interval (95% CI), and P-values (P) estimated with Cox regression testing for an association between the p.G137S variant and risk of different types of CVD events.

**Table S5. Association between the p.G137S variant and cardiovascular disease outcomes, using a logistic mixed model (GMMAT)**

| Trait | n | Number of events | OR | 95% CI | P |
|---|---|---|---|---|---|
| Any CVD event | 4565 | 627 | 1.17 | 0.97 - 1.42 | 0.10 |
| Ischemic heart disease | 4565 | 237 | 1.50 | 1.15 - 1.97 | 0.0029 |
| Cerebrovascular disease | 4565 | 362 | 1.06 | 0.85 - 1.34 | 0.60 |
| Peripheral artery disease | 4565 | 47 | 1.69 | 1.00 - 2.84 | 0.050 |
| Heart failure | 4565 | 161 | 1.11 | 0.79 - 1.55 | 0.54 |
| Coronary operations | 4565 | 88 | 1.75 | 1.16 - 2.65 | 0.0080 |

Odds ratio (OR), 95% confidence interval (95% CI), and P-values (P) estimated with a GLMM testing for an association between the p.G137S variant and risk of different types of CVD events, using GMMAT.

### Table S6. Hazard ratio for selected CVD events with and without adjustment for circulating lipid concentrations

| Adjustment | n | Ischaemic heart disease | Peripheral artery disease | Heart failure | Coronary operations |
|---|---|---|---|---|---|
| None | 4565 | 1.51 (1.18-1.92) | 1.69 (1.01 - 2.82) | 1.07 (0.78 - 1.46) | 1.78 (1.21 - 2.62) |
| LDL cholesterol | 3851 | 1.32 (0.97-1.80) | 2.25 (1.20-4.21) | 1.20 (0.83-1.74) | 1.71 (1.07-2.72) |
| HDL cholesterol | 4543 | 1.47 (1.15-1.87) | 1.88 (1.12-3.16) | 1.05 (0.76-1.43) | 1.73 (1.17-2.55) |
| Total cholesterol | 4409 | 1.41 (1.08-1.86) | 1.93 (1.10-3.39) | 1.32 (0.81-1.61) | 1.49 (0.97-2.31) |
| Apolipoprotein B | 1232 | 1.57 (1.05-2.33) | 1.51 (0.64-3.6) | 1.28 (0.78-2.09) | 2.44 (1.33-4.45) |

Risk estimates (hazard ratios (95% confidence intervals)) from a Cox regression testing for an association between the p.G137S variant and four types of CVD events. The analyses were adjusted for raw values of LDL cholesterol, HDL cholesterol, total cholesterol, and apolipoprotein B, respectively, and run on all available data for each trait.

### Table S7. Classification codes for cardiovascular disease events

| | ICD8 | ICD10 | ICPC2 |
|---|---|---|---|
| Ischemic heart disease | 41199, 41009, 41099, 41109, 41209, 41299, 41409, 41499 | DI21, DI22, DI23, DI24, DI25 | K75, K76 |
| Cerebrovascular disease | 43100, 43101, 43108, 43109, 43190, 43191, 43198, 43199, 43200, 43201, 43202, 43208, 43209, 43290, 43291, 43292, 43293, 43298, 43299, 43309, 43399, 43409, 43499, 43599, 43509, 43601, 43609, 43690, 43699, 43700, 43701, 43708, 43709, 43790, 43791, 43798, 43799 | DI61, DI62, DI63, DI64, DI65, DI66, DI672, DI678, DI693, DI694, DG458, DG459 | K89, K90, K91 |
| Peripheral artery disease | 44009, 44019, 44020, 44021, 44028, 44029, | DI70, DI739, DI739A, DI739C, DI740, DI740B, | K92, K99 |

| | | | |
|---|---|---|---|
| | 44099, 44039, 44408, 44409, 44419, 44420, 44421, 44428, 44442, 44429, 44439, 44440, 44441, 44499, 44490, 44449, 44448, 44443, 44444 | DI740D, DI741, DI742, DI743, DI744, DI745, DI748, DI749 | |
| Heart failure | 78249, 42719, 42710, 42711, 42899, 42709 | DI50 | K77 |

ICD, International Statistical Classification of Diseases and Related Health Problems; ICPC, International Classification of Primary Care.

**Table S8. Classification codes for operational procedures related to cardiovascular health**

| | **The Classification of Operations and Treatments** | **NOMESCO Classification of Surgical Procedures** |
|---|---|---|
| Coronary operations | 30359, 30354, 30350, 30245, 30241, 30240, 30200, 30199, 30189, 30179, 30169, 30159, 30149, 30139, 30129, 30120, 30119, 30109, 30099, 30089, 30079, 30069, 30059, 30049, 30039, 30029, 30009, 30019 | KFNA, KFNB, KFNC, KFND, KFNE, KFNF, KFNG, KFNH, KFNW96, KFNW98 |

NOMESCO, Nordic Medico-Statistical Committee.
